# Sleep Quantity and Quality During Inpatient Rehabilitation After Stroke

**DOI:** 10.1101/2023.03.22.23287526

**Authors:** George Fulk, Sandra Billinger, Bria Bartsch, Pamela Duncan, Daniel Valastro, Karen Klingman

## Abstract

**Objective:** To identify sleep patterns and their association with recovery after stroke during inpatient rehabilitation, and to determine if clinical outcomes are different between participants demonstrating abnormal sleep patterns as compared to those with normal sleep patterns.

**Methods:** Cohort study in which participants were undergoing inpatient rehabilitation after a stroke. Sleep quantity and quality was measured using an actigraph that participants wore for up to 7 nights during the first week of inpatient rehabilitation. Medicare Quality Indicators (GG code), Barthel Index, gait speed, and Berg balance scale were collected at admission and discharge. Participants were categorized into groups based on meeting or not meeting recommended sleep quantity and quality guidelines. Association between sleep patterns and outcomes were assessed using Pearson correlation and differences in outcomes and length of stay between participants who met or did not meet sleep quantity and quality guidelines were determined using independent sample t-test.

**Results:** 69 participants were in the study. Sleep quantity and quality was poor for all the participants. None of the participants met all the sleep quantity and quality guidelines. There were moderate to small associations (-0.42 to 0.22) between some sleep quantity and quality parameters and clinical outcomes. Participants who’s sleep efficiency (SE) was <85% had a significantly longer length of stay compared to those who’s SE was >=85% (17.4 vs. 21.5 days, p<0.05).

**Conclusions:** People with stroke undergoing inpatient rehabilitation have poor sleep quantity and quality. There is a small to moderate association between sleep patterns and clinical outcomes and participants with poor sleep quality had longer length of stay compared to those with good sleep quality. Further research is necessary to better understand the complex relationship between sleep and recovery after stroke.

**Impact:** Sleep is associated with functional recovery during inpatient rehabilitation after stroke.

## Introduction

Sleep is vital for health and quality of life. Although sleep disorders are not fully understood in people with stroke, sleep disruption in people with stroke is likely multifactorial and negatively impacts recovery. Some sleep disorders may be preexisting, for example obstructive sleep apnea is a risk factor for stroke^1^ and may be a result after stroke.^1,2^ Other factors such as lesion location, depression, fatigue, limited mobility in bed, and the hospital environment may contribute to disrupted sleep.^3^ Evidence is growing that indicates that sleep plays an important role during recovery after stroke, with sleep directly impacting the critical rehabilitation component of motor relearning following stroke In people with chronic stroke, Siengsukon and colleagues^4^ found that greater sleep efficiency, less time spent in stage NREM3 sleep, and more time spent in stage REM sleep was associated with the magnitude of motor learning on an upper extremity tracking task.

Sleep disorders and poor sleep may also play a role in recovery during the acute and subacute stages of stroke. A recent systematic review reported that people with stroke with obstructive sleep apnea (OSA) had poorer function and greater disability than those with stroke without OSA. The impact of other sleep disorders on recovery was less clear, possibly due to the smaller number of studies and lack of consistent methodology for diagnosing other sleep disorders.^5^ Poor sleep itself, regardless of a specific diagnosed sleep disorder, early after stroke may also negatively impact recovery. Iddagoda and colleagues^6^ found that poor self-reported sleep quality using the Pittsburgh Sleep Quality Index (PSQI) was related to less change on the Functional Independence Measure (FIM) during inpatient rehabilitation. Chow and colleagues^7^ found that total sleep time, measured using accelerometry, was negatively associated with time in sedentary activity and positively associated with time in light and moderate activity, measured using accelerometry, during inpatient rehabilitation. Poor sleep may also impact other aspects of health in addition to function after stroke. Self-reported poor sleep 2 weeks after stroke may be associated with depression and anxiety 3 months later.^8^

The impact of sleep on recovery after stroke is not fully understood. There is a need to better understand sleep patterns of people with stroke, the potential associations between sleep and demographic and clinical outcomes, and if people with stroke who have abnormal sleep patterns have poorer outcomes during inpatient rehabilitation in order to develop personalized interventions to improve sleep and enhance recovery during inpatient rehabilitation. Previous studies have used a mix of objective and subjective measures of sleep and have not considered pre-existing OSA, and none have examined if people with stroke who have abnormal sleep patterns have poorer clinical outcomes. The purposes of this study were: 1) to report on the sleep patterns of people with stroke without OSA during the first week of inpatient rehabilitation and to determine the number of participants who demonstrated abnormal sleep patterns, 2) to explore associations between sleep patterns, demographics and clinical outcomes, and 3) to determine if clinical outcomes were different between participants demonstrating abnormal sleep patterns as compared to those with normal sleep patterns.

## Methods

Participants were recruited from two inpatient rehabilitation hospitals, one in the Northeast and one in the Midwest of the United States, and are part of a larger, ongoing cohort study to determine the prevalence of non OSA sleep disorders and their impact on recovery after stroke.^9^ Inclusion criteria were: diagnosis of stroke by neuroimaging determined by review of the medical record, >= 18 years old, National Institutes of Health Stroke Scale (NIHSS) item 1a score <2 (alert or not alert but arousable by minor stimulation to obey, answer, or respond), provide informed consent or assent, with the participant’s legal guardian providing consent. Exclusion criteria were: pre-stroke diagnosis of OSA by review of medical history, oxygen desaturation index (ODI) >=15, living in nursing home or assisted living center prior to stroke, unable to ambulate at least 150’ independently prior to stroke, other neurologic health condition such as Parkinson disease or Multiple Sclerosis that could impact recovery, pregnant, recent hemicraniectomy or suboccipital craniectomy, planned discharge >150 miles from recruiting hospital, and global aphasia as determined by a score of 3 on NIHSS item 9. The funders played no role in the design, conduct, or reporting of this study. All participants provided consent or assent (with participant’s legal guardian providing consent) and the study was approved by WCG IRB (20202548).

Demographic variables were collected through patient interview and review of the medical record and include age, sex, race, ethnicity, type of stroke (ischemic or hemorrhagic), location of stroke (left hemisphere, right hemisphere, left subcortical, right subcortical, brainstem, cerebellar, bilateral), smoking status (current, smoked in the past prior to current stroke but quit at least 3 months before admitted to the hospital, never), and marital status (single, married, separated, divorced, widowed, other).

The following clinical outcomes were collected through a combination of review of the medical record and interview with the participant: National Institutes of Health Stroke Scale (NIHSS)^10^ to measure stroke severity; self-care, mobility and total Section GG (SC-GG, Mob-GG, Total-GG) assessment scores which are utilized by the Centers for Medicare and Medicaid Services (at admission and discharge) to measure overall functional ability; Barthel Index^11^ (at admission and discharge, BI) to measure overall functional ability; Berg Balance Scale^12^ (at admission and discharge, BBS) to measure functional balance; gait speed measured over the middle six meters of a ten-meter walk^13^ (at admission and discharge, GS) to measure walking ability; Montreal Cognitive Assessment (MoCA)^14^ to measure cognition; and Patient Health Questionnaire 9 (PHQ-9)^15^ to measure depression. We also calculated GG code efficiency for SC-GG, Mob-GG, and Total-GG, discharge score – admission score divided by length of stay (LOS) (SC-GG-E, Mob-GG-E, and Total-GG-E). Additionally, LOS in inpatient rehabilitation was collected.

After providing informed consent, or assent, the participant wore a wrist nocturnal oximeter (Nonin Model 3150) for one night to determine if their ODI was less than 15. If it was, the participant continued in the study. Sleep patterns, total sleep time (TST), sleep efficiency (SE), wake after sleep onset (WASO), number of awakenings (NWAK) per night, and mean duration of awakenings (DA) were measured using a wGT3X-BT Actigraph (actigraph).^16-19^ The actigraph sleep watch (https://actigraphcorp.com/actigraph-wgt3x-bt/), with non-wear detection and ambient light sensing, was worn on the participant’s least affected wrist for 24 hours per day for up to 7 days during the first week of inpatient rehabilitation. Actigraphs were set to collect data at 30Hz sample rate, and later uploaded for analysis as 60 second epochs with three-axis motion detection. At the conclusion of each timepoint’s wear period, watch data were transferred from the actigraph to a study computer for analysis using ActiGraph’s ActiLife v6.13.4 software. Wear times were validated (marked as “wear” if either Troiano method or ActiGraph sensor indicated “wear”), and sleep parameters were determined via the Cole Kripke algorithm and Actigraph sleep period detection method. First-pass software-calculations of time-to-bed and time-out-of-bed were sometimes adjusted prior to calculation of sleep outcome variables, under certain conditions. For overnight timeframes with two separate sleep periods detected by the software, a single sleep period spanning the combination of both originally detected periods was substituted. If the ActiLife software was unable to determine time to bed and time out of bed, every effort was made to manually determine these from activity and light signals in the ActiLife interface. Any sleep periods appearing to be daytime naps were not included in analysis of nightly sleep.

Participants were dichotomized into sleep pattern groups based on recommendations from the American Academy of Sleep Medicine, Sleep Research Society, and National Sleep Foundation^20,21^ according to average TST < than seven hours/night vs. ≥ seven hours/night (-TST vs. +TST), average SE < 85% vs. ≥ 85% (-SE vs. +SE), average WASO ≥31 minutes/night vs. <31 (-WASO vs. +WASO), and average NWAK/night >3 vs. ≤3 (-NWAK vs. +NWAK).

## Data Analysis

R, version 4.2.1, was used for data analysis. Data from participants were used only if they had at least one full night of sleep data. Descriptive statistics (mean and standard deviation) were used to describe the mean sleep patterns per night (TST, SE, WASO, NA, and mean DA) of people with stroke without OSA, and the number of participants who demonstrated abnormal sleep patterns were identified (first purpose).

Pearson correlation coefficients were used to explore the association between sleep patterns (TST, SE, NA, mean DA) and clinical outcomes (NIHSS, SC-GG, Mob-GG, Total-GG, BI, BBS, GS, MOCA, PHQ-9, SC-GG-E, Mob-GG-E, Total-GG-E, and LOS) (second purpose).

Independent t-tests were used to determine if there were differences in clinical outcomes (SC-GG, Mob-GG, Total-GG, BI, BBS, GS, MOCA, PHQ-9, SC-GG-E, Mob-GG-E, Total-GG-E, and LOS) between sleep pattern groups (-TST vs. +TST, -SE vs. +SE, -WASO vs. +WASO, and -NWAK vs. +NWAK) (third purpose). Because these data were collected as part of a larger ongoing study, we did not determine sample size to detect differences between these groups a priori and due to the exploratory nature of the study we did not correct the p value for multiple comparisons. P was set to < 0.05 for significance for all analyses.

## Results

Sixty-nine people with stroke without OSA participated in the study. There were 29 men and 40 women, who were admitted to inpatient rehabilitation a mean 9.9 (8.0) days after their stroke.Participants presented with a mild to moderate stroke, mean NIHSS 5.6 (3.6), and were moderately limited in activities of daily living, mean admission BI 39.6 (22.6). See Table 1 for demographics and clinical outcomes.

**Table 1.**
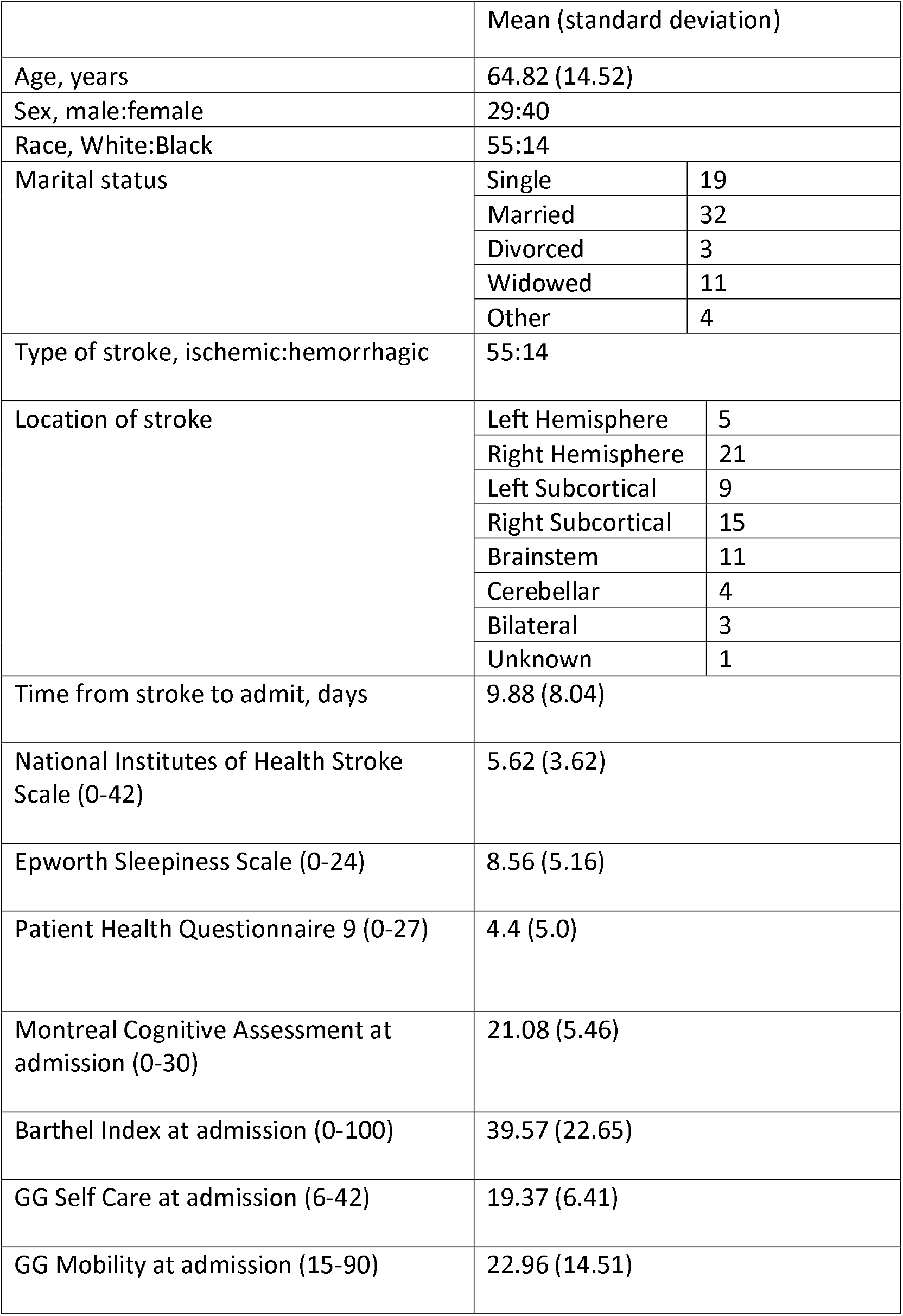
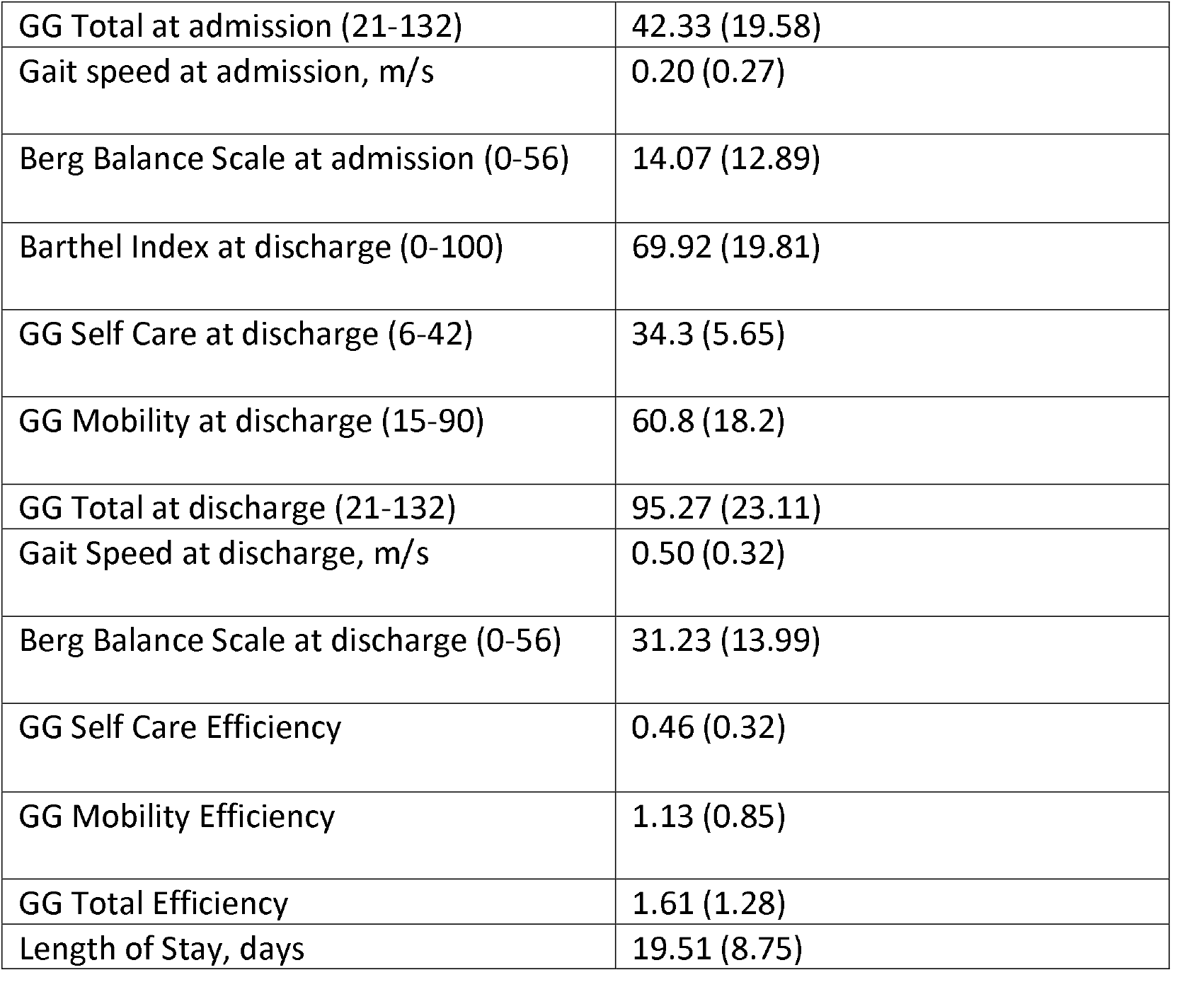
Demographic and Clinical Outcomes

|  | Mean (standard deviation) |  |
| --- | --- | --- |
| Age, years | 64.82 (14.52) |  |
| Sex, male:female | 29:40 |  |
| Race, White:Black | 55:14 |  |
| Marital status | Single | 19 |
|  | Married | 32 |
|  | Divorced | 3 |
|  | Widowed | 11 |
|  | Other | 4 |
| Type of stroke, ischemic:hemorrhagic | 55:14 |  |
| Location of stroke | Left Hemisphere | 5 |
|  | Right Hemisphere | 21 |
|  | Left Subcortical | 9 |
|  | Right Subcortical | 15 |
|  | Brainstem | 11 |
|  | Cerebellar | 4 |
|  | Bilateral | 3 |
|  | Unknown | 1 |
| Time from stroke to admit, days | 9.88 (8.04) |  |
| National Institutes of Health Stroke Scale (0-42) | 5.62 (3.62) |  |
| Epworth Sleepiness Scale (0-24) | 8.56 (5.16) |  |
| Patient Health Questionnaire 9 (0-27) | 4.4 (5.0) |  |
| Montreal Cognitive Assessment at admission (0-30) | 21.08 (5.46) |  |
| Barthel Index at admission (0-100) | 39.57 (22.65) |  |
| GG Self Care at admission (6-42) | 19.37 (6.41) |  |
| GG Mobility at admission (15-90) | 22.96 (14.51) |  |

|  |  |
| --- | --- |
| GG Total at admission (21-132) | 42.33 (19.58) |
| Gait speed at admission, m/s | 0.20 (0.27) |
| Berg Balance Scale at admission (0-56) | 14.07 (12.89) |
| Barthel Index at discharge (0-100) | 69.92 (19.81) |
| GG Self Care at discharge (6-42) | 34.3 (5.65) |
| GG Mobility at discharge (15-90) | 60.8 (18.2) |
| GG Total at discharge (21-132) | 95.27 (23.11) |
| Gait Speed at discharge, m/s | 0.50 (0.32) |
| Berg Balance Scale at discharge (0-56) | 31.23 (13.99) |
| GG Self Care Efficiency | 0.46 (0.32) |
| GG Mobility Efficiency | 1.13 (0.85) |
| GG Total Efficiency | 1.61 (1.28) |
| Length of Stay, days | 19.51 (8.75) |

Participants wore the actigraph for a mean of 4.4 (1.7) nights. The mean TST and of all the sleep patterns was below that recommended by the American Academy of Sleep Medicine, Sleep Research Society, and National Sleep Foundation, see Table 2 and Figure 1a-d. Sixty two point three percent (62.3%, 43/69) of the participants got less than 7 hours of sleep per night on average, 50.7% (35/69) of the participants’ SE was less than 85% on average, 87.0% (60/69) of the participants were awake after initially falling asleep for more than 30 minutes on average, and 98.6% (68/69) of the participants woke up more than 3 times during the night on average. None of the participants met recommended cut offs indicating good sleep amount or good sleep quality across TST, SE, WASO, and NWAK. Only four participants met recommended cut offs indicating sufficient sleep amount and good sleep quality across 3 of the 4 sleep patterns, see Table 3.

**Table 2.** Mean Sleep Quantity and Quality During the First Week of Inpatient Rehabilitation

| Sleep Pattern | Mean (standard deviation) |
| --- | --- |
| Total Sleep Time, minutes | 392.34 (91.48) |
| Sleep Efficiency, % | 83.03 (8.98) |
| Wake After Sleep Onset, minutes | 82.47 (50.16) |
| Number of Awakenings | 13.12 (6.25) |
| Mean Duration of Awake Episode, minutes | 6.42 (3.36) |

**Table 3.** Number of Participants Categorized with Poor Sleep Quantity and Quality Across Different Sleep Patterns

| Total Sleep Time | Sleep Efficiency | Wake After Sleep Onset | Number of Awakenings | Number of Participants |
| --- | --- | --- | --- | --- |
| < 7 hours | < 85% | > 30 minutes | > 3 | 25 |
| < 7 hours | >=85% | > 30 minutes | > 3 | 12 |
| < 7 hours | >=85% | <=30 minutes | > 3 | 5 |
| < 7 hours | >=85% | <=30 minutes | <= 3 | 1 |
| >= 7 hours | < 85% | > 30 minutes | > 3 | 10 |
| >= 7 hours | >=85% | > 30 minutes | > 3 | 13 |
| >= 7 hours | >=85% | <=30 minutes | > 3 | 3 |

**Figure 1a.**
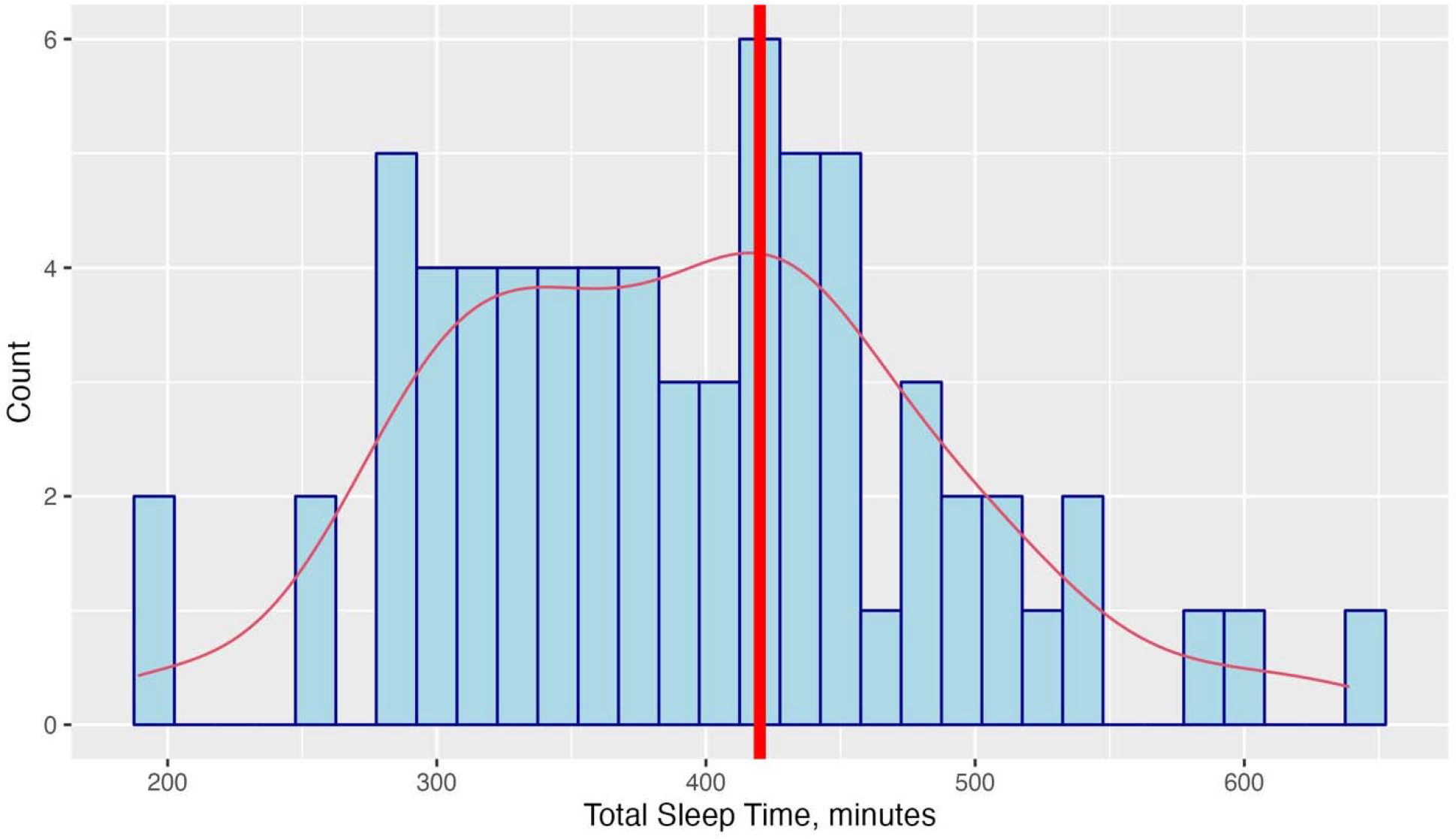
Distribution of Total Sleep Time.

**Figure 1b.**
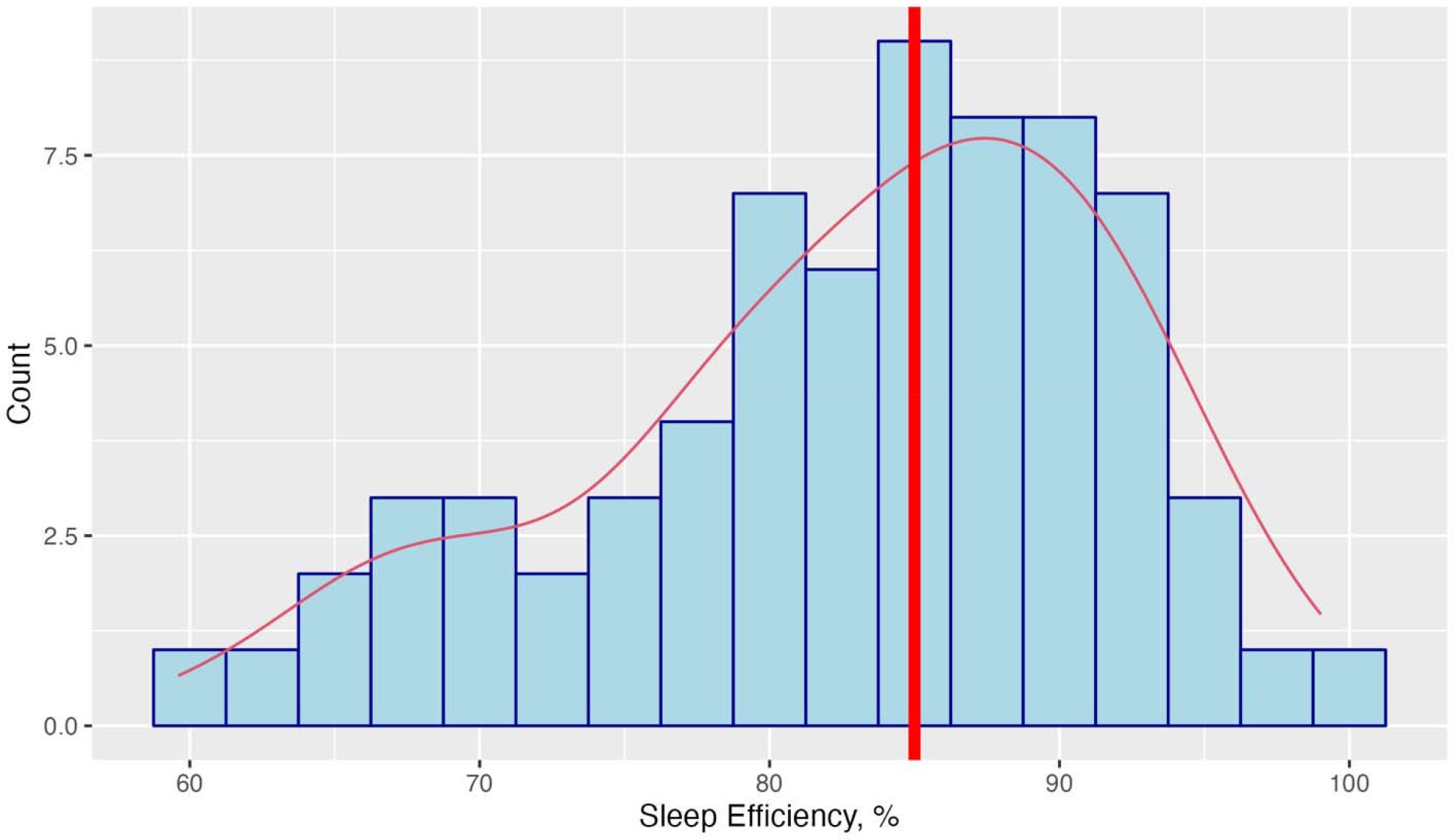
Distribution of Sleep Efficiency.

**Figure 1c.**
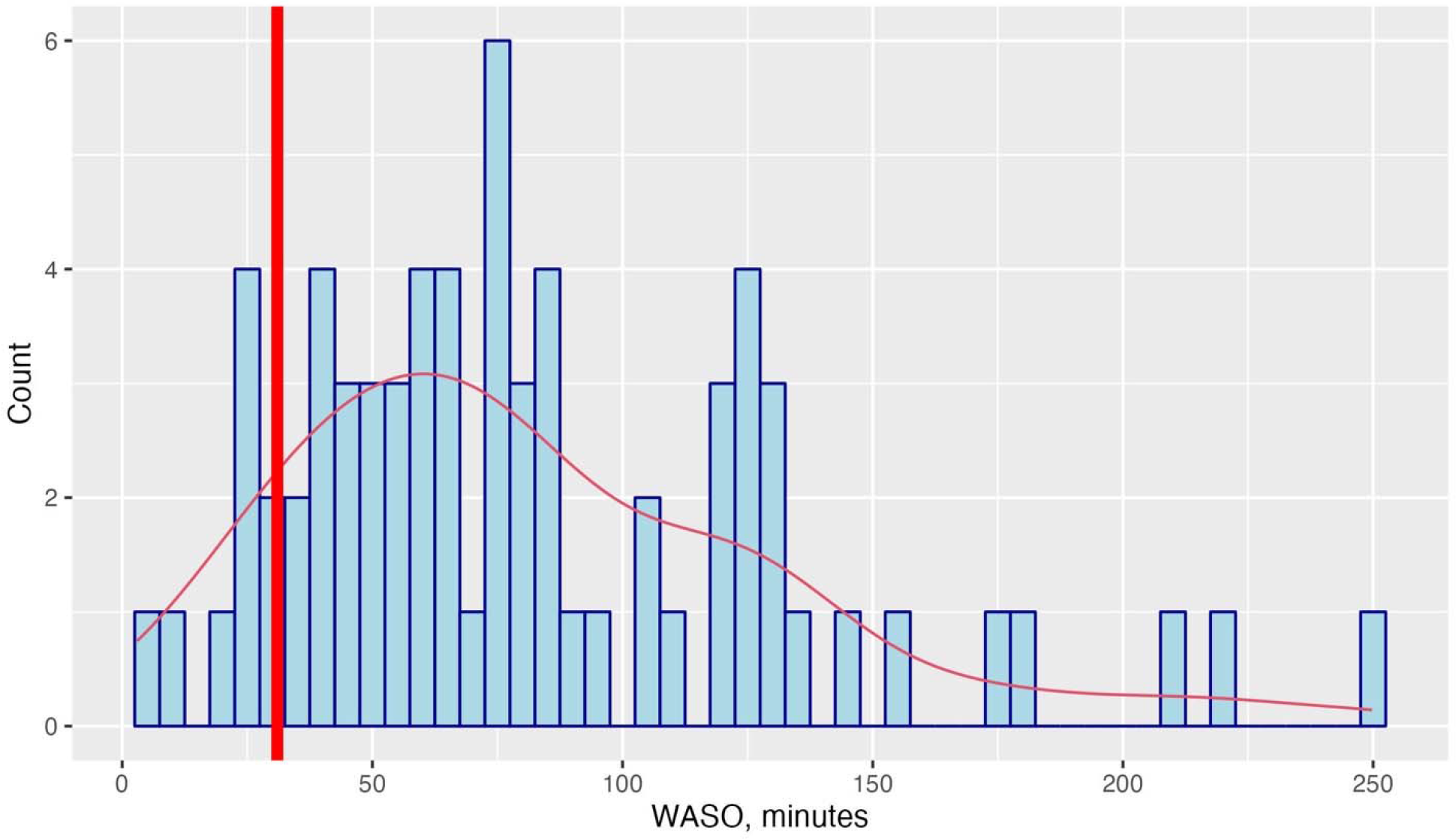
Distribution of Wake After Sleep Onset.

**Figure 1d.**
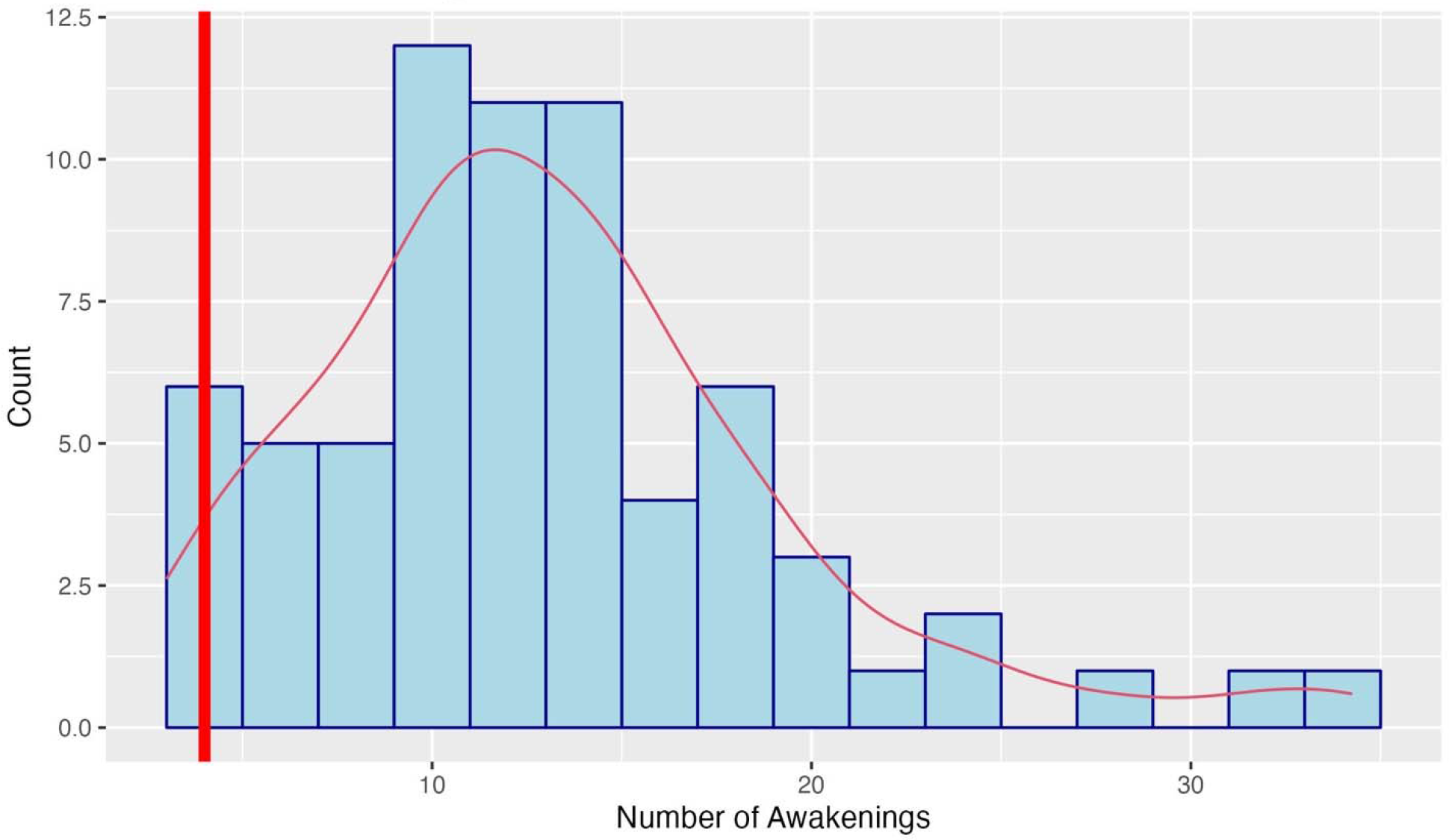
Number of Awakenings.

There was no association between sleep patterns and most of the demographic and clinical outcomes except for the following: mean DA was significantly associated with NIHSS (r=0.24 [p=0.05]) and LOS (r=0.35 [0.009]); SE was significantly associated with LOS (r=-0.42 [p=0.01]), SC-GG-E (r=0.30 [p=0.015]), Mob-GG-E (r=0.28 [p=0.03]), and Total-GG-E (r=0.29 [p=0.01]), see Table 4 and Figure 2a-d. The associations between TST and SC-GG-E, Mob-GG-E, Total-GG-E, GS at discharge; WASO and SC-GG-E; and DA and Mob-GG-E and Total-GG-E approached significance and were in the direction of poor sleep indicating worse outcomes, see Table 4.

**Table 4.** Associations Between Sleep Quantity, Sleep Quality, and Clinical Outcomes

|  | Total Sleep Time | Sleep Efficiency | Wake After Sleep Onset | Mean Duration of Awakenings |
| --- | --- | --- | --- | --- |
| National Institutes of Health Stroke Scale |  |  |  | 0.24 (0.04) |
| Length of Stay |  | -0.42 (0.01) |  | 0.35 (0.009) |
| GG Self Care Efficiency | 0.22 (0.08) | 0.29 (0.02) | -0.24 (0.059) |  |
| GG Mobility Efficiency | 0.23 (0.07) | 0.28 (0.03) |  | -0.24 (0.059) |
| GG Total Efficiency | 0.25 (0.052) | 0.29 (0.02) |  | -0.25 (0.052) |
| Discharge Gait Speed | 0.26 (0.057) |  |  |  |

**Figure 2a.**
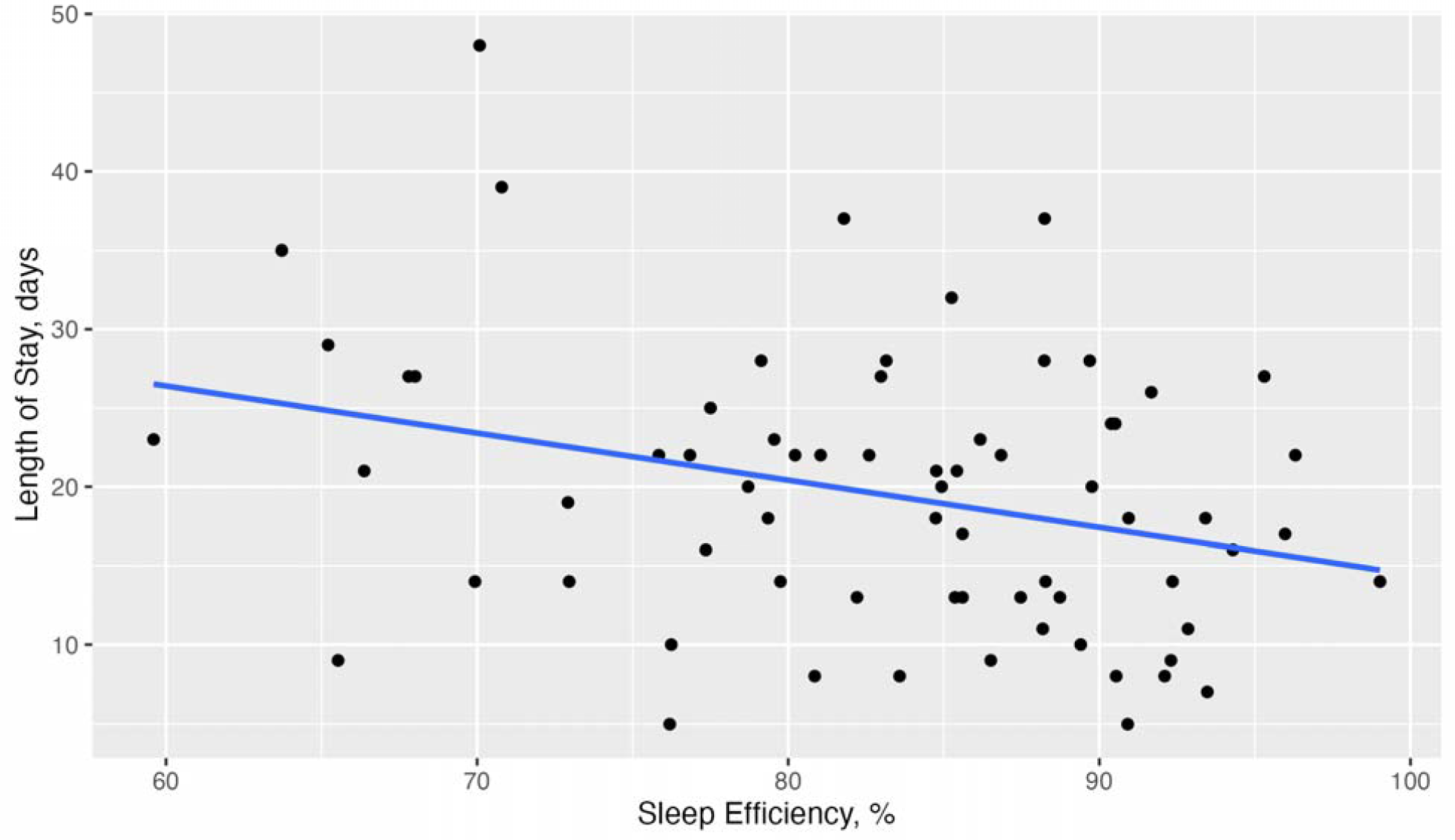
Sleep Efficiency and Length of Stay.

**Figure 2b.**
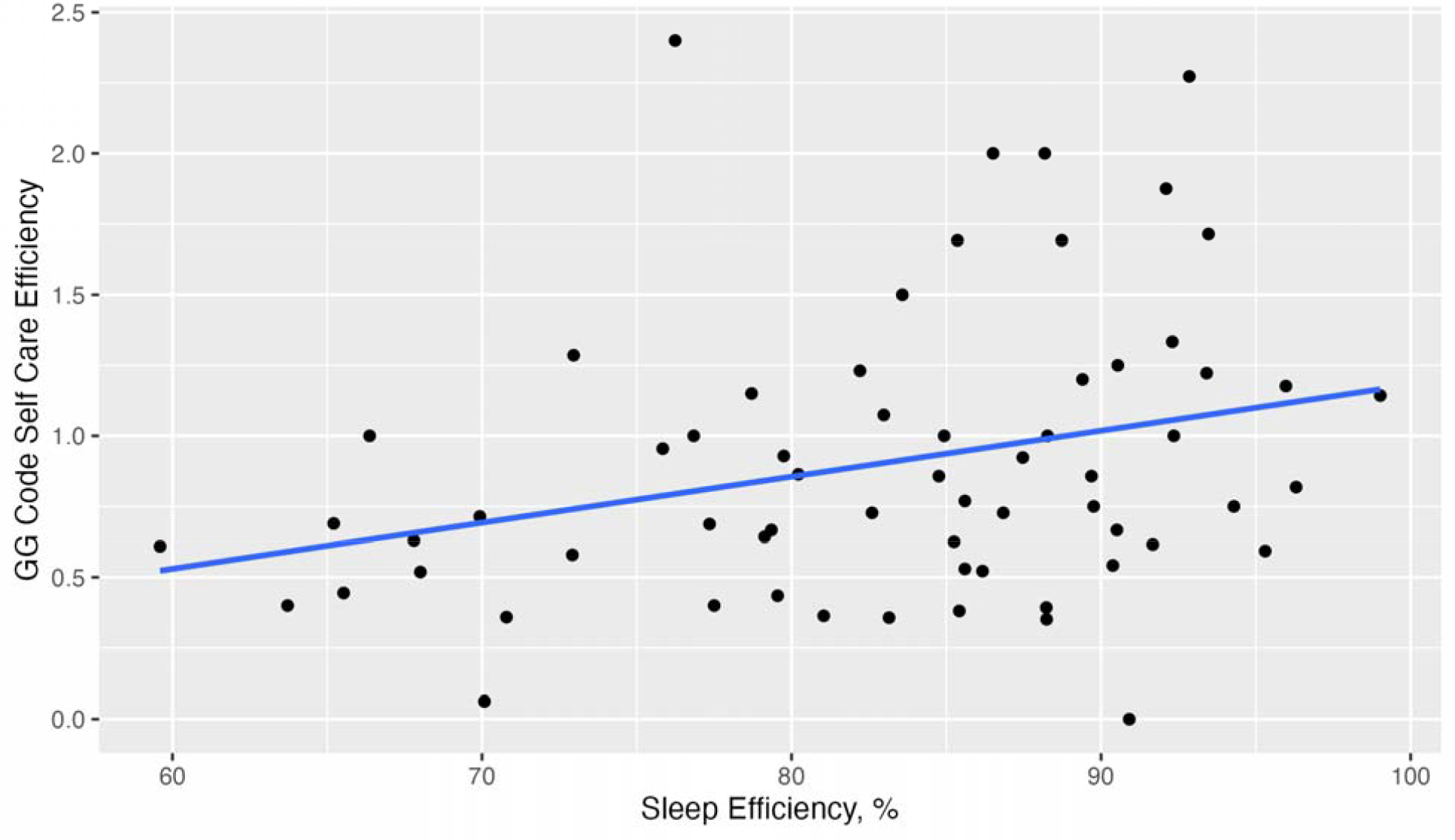
Association Between Sleep Efficiency and GG Code Self Care Efficiency.

**Figure 2c.**
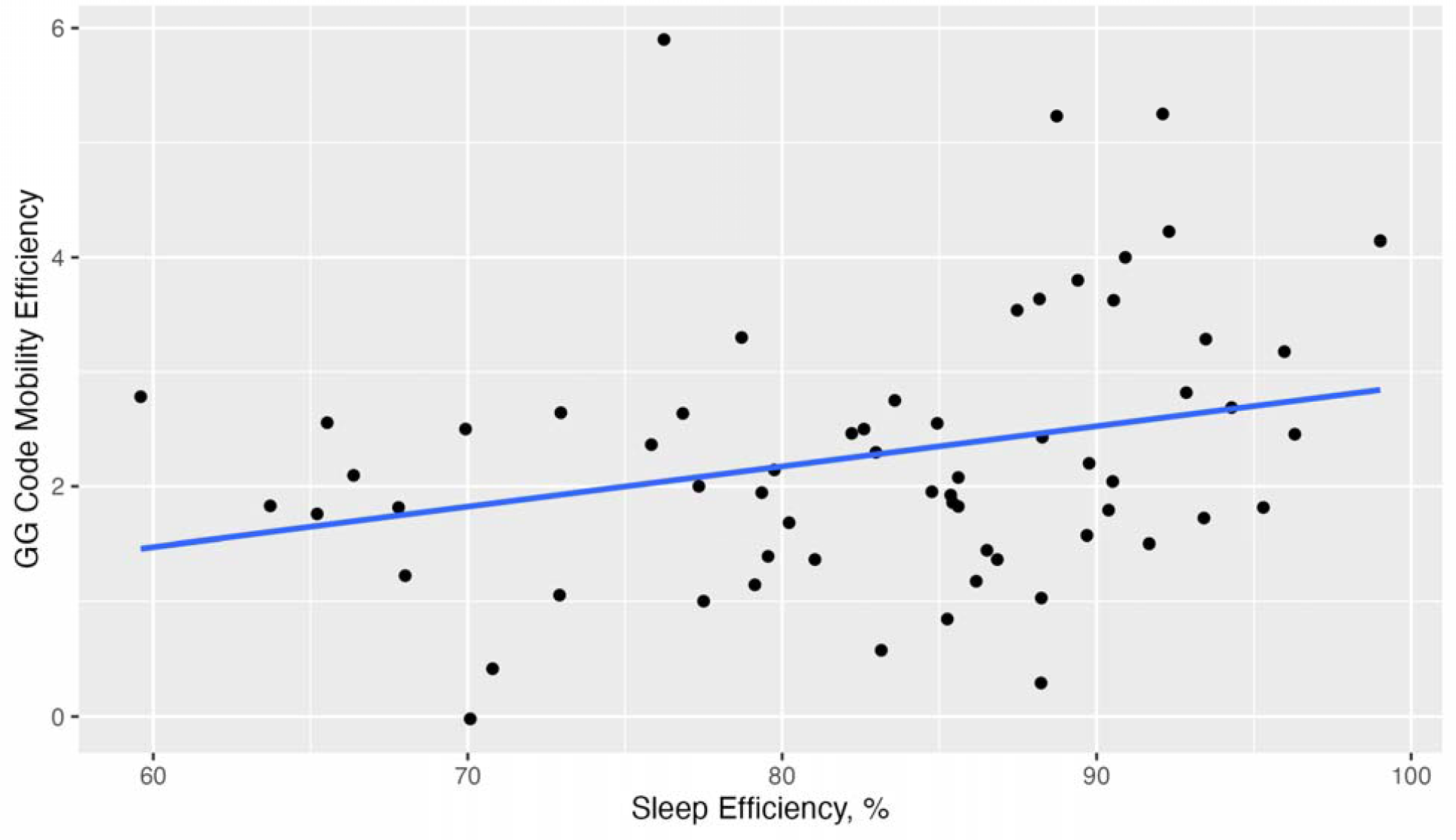
Association Between Sleep Efficiency and GG Code Mobility Efficiency.

**Figure 2d.**
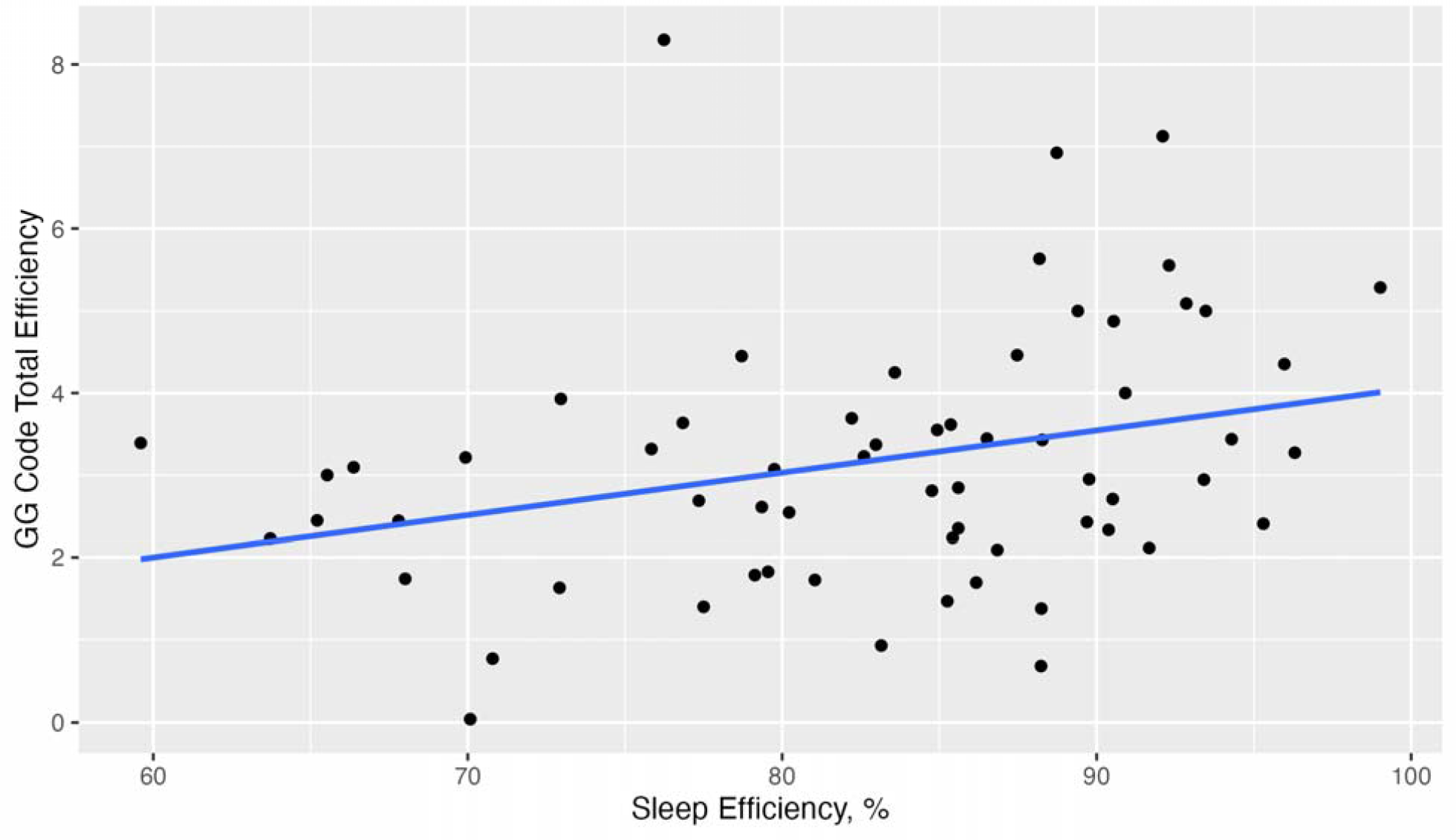
Association Between Sleep Efficiency and GG Code Total Efficiency.

Because there was a large percentage of participants who did not meet recommended WASO and NWAK guidelines, 87.0% and 98.6% respectively, we did not perform t-tests comparing clinical outcomes between groups for those two sleep pattern criteria. An independent t-test comparing the mean length of stay between the -SE group (SE <85%) and the +SE group (SE >=85%) found a significant difference between the mean LOS between the two groups, p=0.049). The mean LOS of the -SE group was significantly longer, 21.5 days, than the mean LOS of the +SE group, 17.4 days, Figure 3. There were no other significant differences in clinical outcomes (SC-GG, Mob-GG, Total-GG, SC-GG-E, Mob-GG-E, Total-GG-E, BI, BBS, GS, MOCA, and PHQ-9) between the different sleep groups (−TST vs. +TST, -SE vs. +SE, -WASO vs. +WASO, and -NWAK vs. +NWAK).

**Figure 3.**
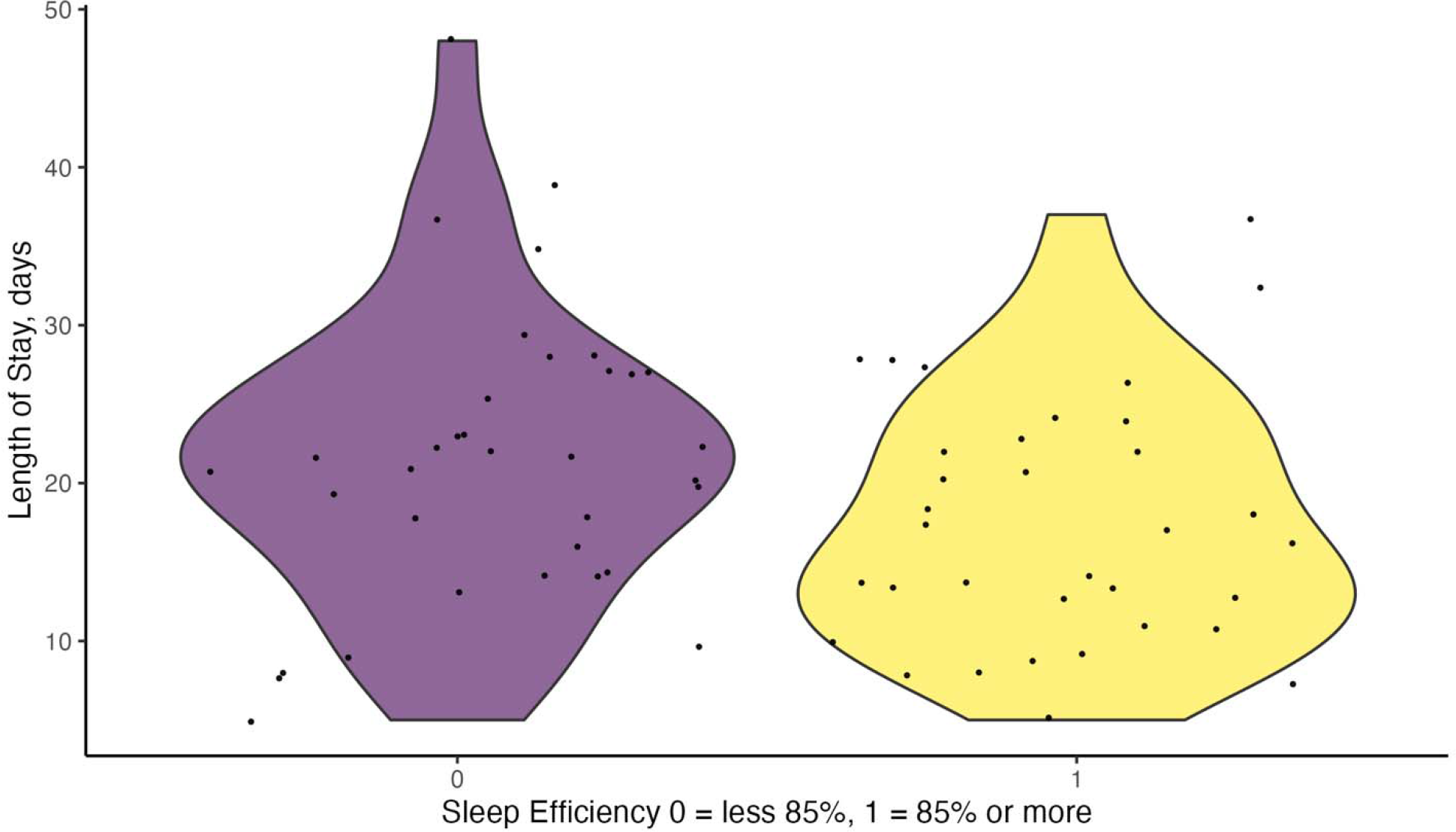
Length of Stay Between Sleep Efficiency Groups.

## Discussion

The quantity and quality of sleep during inpatient rehabilitation was poor for all participants. No participant met all the recommendations for quantity of sleep (TST >=7 hours) and quality of sleep, SE (>=85%), time awake after initially falling asleep (<=30 minutes), or NWAK (<=3) during the first week of inpatient rehabilitation. Only four participants met three of the four sleep quantity and quality recommendations, while 36% (25/69) did not meet any of the sleep recommendations.

Although alarming, these findings of poor sleep quantity and quality are not surprising given the challenges with sleeping in the hospital. Hospitalized patients often report insufficient and disturbed sleep.^22,23^ Abnormal sleep in the hospital can lead to long- and short-term negative consequences. In the short-term, disrupted sleep in the hospital can lead to poor glycemic control, pain intensity, reduced respiratory function, and impaired muscle strength.^23^ In the long term after discharge, disrupted sleep during a stay in the hospital can lead to chronic pain,^24^ chronic insomnia,^25^ and higher mortality^26^.

There are some similarities and differences in the sleep quantity and quality of the participants in this study compared to other studies. The mean TST (392 minutes, 62% slept less than 7 hours) was less than most other studies^27,28,29,7^ which all reported TST means of their samples above 420 minutes or more than 50% of the sample having a TST >=420 minutes. Although the mean SE (83% and more than half of the participants exhibiting SE less than 85%) was less than recommended, it was slightly higher than the SE reported by Kim and colleagues^29^ (77%). The NWAK (13) in our sample was the same as those reported by Bakken and colleagues^27^ but less than those reported by Kim and colleagues.^29^ The WASO in our study was similar to others.^29,30^ Interpreting these differences and similarities is challenging. None of these other studies reported the full breadth of objectively measured sleep quantity and quality measures that we did. Even though all studies measured sleep while participants were in the hospital, the length of time since stroke to when sleep was measured was different, with the length of time in our study less than the others. Importantly, our findings provided broader evidence of disrupted sleep time and quality in people with stroke when they are participating in inpatient rehabilitation.

Although some of the associations between sleep quantity/sleep quality and clinical outcomes were relatively small, we believe these may still be clinical important. During this early stage of recovery there are many factors such as motor function,^31,32^ balance,^33^ age, time from stroke to admission to inpatient rehab, BMI, and race that impact recovery.^34^ Our finding that sleep during the first week of inpatient rehabilitation is associated with functional outcomes at discharge, approximately 20 days after admission, perhaps indicates that sleep plays a vital role during the recovery process and should be investigate further.

Similar to our findings, other studies in stroke have found associations between certain sleep patterns and clinical outcomes and no associations between others. For example, Bakken and colleagues^27^ found a small association between TST and WASO and activities of daily living, while Wiliams-Cooke^28^ and colleagues found no association between TST and function. We believe these different findings illustrate the need for further research to better understand the mechanisms of the impact of sleep on recovery after stroke.

Unique in our study was the impact of SE during the first week of inpatient rehabilitation on outcomes. SE was inversely associated with LOS, where participants with poorer SE had longer lengths of stay, and was positively associated with efficiency of functional recovery, where participants with greater SE had greater functional recovery in less time. Additionally, participants who did not meet the recommended SE for healthy sleep had a statistically significant longer LOS compared to those who had a SE >=85%, which is recommended as an indication of good sleep quality. This finding supports the importance of good quality sleep during recovery.

The impact of sleep quality on recovery could be multifactorial. Siegnsukon and colleagues^35-37^ have found that sleep, in people with stroke, modulated motor learning, and they postulated that motor recovery after stroke may be modulated by sleep. We found that sleep also impacts overall recovery during hospitalization such that the participants with higher quality sleep recovered more quickly and efficiently. Enhancing the hospital environment to help facilitate better sleep quality may help improve recovery during inpatient rehabilitation. Lighting could be improved, setting specific quiet times during the night, and reducing the disruptions such as taking vital signs during the early morning hours should be explored as ways to improve recovery in the hospital.^38^

## Limitations

This study has some limitations that impact the interpretation of the findings. Primarily, we only measured sleep during the first week of inpatient rehabilitation, not throughout the entire length of stay. We don’t know how sleep may have changed throughout the course of rehabilitation and how this may have impacted recovery. We also estimated sleep patterns from actigraphy, not the gold standard for measuring sleep, polysomnography. Polysomnography may have provided greater insight into the impact and mechanisms of sleep on recovery. Although we had data on general location of stroke, we did not have specific stroke location. These data may also have provided more insight into mechanisms of the impact of sleep.

Sleep quantity and quality are disrupted during inpatient rehabilitation in people with stroke. Poor sleep quality is associated with less efficient and longer recovery. These findings can help guide the development of interventions to enhance sleep which in turn may improve recovery after stroke. Further research is needed to better understand the mechanisms by which sleep impacts recovery after stroke.

## Data Availability

All data produced in the present study are available upon reasonable request to the authors

## Acknowledgements

The authors acknowledge the support of the people with stroke and their care partners who participated in the study. The authors also thank the other members of the research team who assisted with data collection.

## Ethics Approval

This study was approved by WCG IRB (20202548), SUNY Upstate Medical University IRB (1643499), and University of Kansas IRB (00148016).

## References

1. Javaheri S, Barbe F, Campos-Rodriguez F, et al. Sleep Apnea: Types, Mechanisms, and Clinical Cardiovascular Consequences. J Am Coll Cardiol. Feb 21 2017;69(7):841–858. doi:10.1016/j.jacc.2016.11.069

2. Xiaolin Gu MM. Risk factors of sleep disorder after stroke: a meta-analysis. Top Stroke Rehabil. Jan 2017;24(1):34–40. doi:10.1080/10749357.2016.1188474

3. Duss SB, Seiler A, Schmidt MH, et al. The role of sleep in recovery following ischemic stroke: A review of human and animal data. Neurobiol Sleep Circadian Rhythms. Jan 2017;2:94–105. doi:10.1016/j.nbscr.2016.11.003

4. Al-Dughmi M, Al-Sharman A, Stevens S, Siengsukon CF. Sleep characteristics of individuals with chronic stroke: a pilot study. Nature and science of sleep. 2015;7(Journal Article):139–145. doi:10.2147/NSS.S83882 [doi]

5. Fulk GD, Boyne P, Hauger M, et al. The Impact of Sleep Disorders on Functional Recovery and Participation Following Stroke: A Systematic Review and Meta-Analysis. Neurorehabil Neural Repair. Nov 2020;34(11):1050–1061. doi:10.1177/1545968320962501

6. Iddagoda MT, Inderjeeth CA, Chan K, Raymond WD. Post-stroke sleep disturbances and rehabilitation outcomes: a prospective cohort study. Intern Med J. Feb 2020;50(2):208–213. doi:10.1111/imj.14372

7. Hei Chow C, Fraysse F, Hillier S. The relationship between sleep and physical activity in an in-patient rehabilitation stroke setting: a cross-sectional study. Top Stroke Rehabil. Nov 29 2021:1–10. doi:10.1080/10749357.2021.2006982

8. Fan XW, Yang Y, Wang S, et al. Impact of Persistent Poor Sleep Quality on Post-Stroke Anxiety and Depression: A National Prospective Clinical Registry Study. Nat Sci Sleep. 2022;14:1125–1135. doi:10.2147/NSS.S357536

9. Klingman KJ, Skufca JD, Duncan PW, Wang D, Fulk GD. Study Protocol: Sleep Effects on Poststroke Rehabilitation Study. Nurs Res. Nov-Dec 01 2022;71(6):483–490. doi:10.1097/nnr.0000000000000611

10. Goldstein LB, Samsa GP. Reliability of the National Institutes of Health Stroke Scale. Extension to non-neurologists in the context of a clinical trial. Stroke. Feb 1997;28(2):307–10.

11. Quinn TJ, Langhorne P, Stott DJ. Barthel index for stroke trials: development, properties, and application. Stroke. Apr 2011;42(4):1146–51. doi:10.1161/STROKEAHA.110.598540

12. Blum L, Korner-Bitensky N. Usefulness of the Berg Balance Scale in stroke rehabilitation: a systematic review. Phys Ther. May 2008;88(5):559–66. doi:ptj.20070205 [pii] 10.2522/ptj.20070205

13. Fulk GD, Echternach JL. Test-retest reliability and minimal detectable change of gait speed in individuals undergoing rehabilitation after stroke. J Neurol Phys Ther. Mar 2008;32(1):8–13. doi:10.1097/NPT0b013e31816593c0

14. Nasreddine ZS, Phillips NA, Bedirian V, et al. The Montreal Cognitive Assessment, MoCA: a brief screening tool for mild cognitive impairment. J Am Geriatr Soc. 2005;53(4):695–9. doi:10.1111/j.1532-5415.2005.53221.x

15. Williams LS, Brizendine EJ, Plue L, et al. Performance of the PHQ-9 as a screening tool for depression after stroke. Stroke. Mar 2005;36(3):635–8. doi:10.1161/01.STR.0000155688.18207.33

16. Bigue JL, Duclos C, Dumont M, et al. Validity of actigraphy for nighttime sleep monitoring in hospitalized patients with traumatic injuries. J Clin Sleep Med. Feb 15 2020;16(2):185–192. doi:10.5664/jcsm.8162

17. Marino M, Li Y, Rueschman MN, et al. Measuring sleep: accuracy, sensitivity, and specificity of wrist actigraphy compared to polysomnography. Sleep. Nov 1 2013;36(11):1747–55. doi:10.5665/sleep.3142

18. Ancoli-Israel S, Martin JL, Blackwell T, et al. The SBSM Guide to Actigraphy Monitoring: Clinical and Research Applications. Behav Sleep Med. 2015;13(1):S4–S38. doi:10.1080/15402002.2015.1046356

19. Smith MT, McCrae CS, Cheung J, et al. Use of Actigraphy for the Evaluation of Sleep Disorders and Circadian Rhythm Sleep-Wake Disorders: An American Academy of Sleep Medicine Clinical Practice Guideline. J Clin Sleep Med. Jul 15 2018;14(7):1231–1237. doi:10.5664/jcsm.7230

20. Consensus Conference P, Watson NF, Badr MS, et al. Recommended Amount of Sleep for a Healthy Adult: A Joint Consensus Statement of the American Academy of Sleep Medicine and Sleep Research Society. J Clin Sleep Med. Jun 15 2015;11(6):591–2. doi:10.5664/jcsm.475821.

21. Ohayon M, Wickwire EM, Hirshkowitz M, et al. National Sleep Foundation’s sleep quality recommendations: first report. Sleep Health. Feb 2017;3(1):6–19. doi:10.1016/j.sleh.2016.11.006

22. Wesselius HM, van den Ende ES, Alsma J, et al. Quality and Quantity of Sleep and Factors Associated With Sleep Disturbance in Hospitalized Patients. JAMA Intern Med. Sep 1 2018;178(9):1201–1208. doi:10.1001/jamainternmed.2018.2669

23. Elliott R, Chawla A, Wormleaton N, Harrington Z. Short-term physical health effects of sleep disruptions attributed to the acute hospital environment: a systematic review. Sleep Health. Aug 2021;7(4):508–518. doi:10.1016/j.sleh.2021.03.001

24. Smith MT, Klick B, Kozachik S, et al. Sleep onset insomnia symptoms during hospitalization for major burn injury predict chronic pain. Pain. Sep 15 2008;138(3):497–506. doi:10.1016/j.pain.2008.01.028

25. Griffiths MF, Peerson A. Risk factors for chronic insomnia following hospitalization. J Adv Nurs. Feb 2005;49(3):245–53. doi:10.1111/j.1365-2648.2004.03283.x

26. Manabe K, Matsui T, Yamaya M, et al. Sleep patterns and mortality among elderly patients in a geriatric hospital. Gerontology. Nov-Dec 2000;46(6):318–22. doi:10.1159/000022184

27. Bakken LN, Kim HS, Finset A, Lerdal A. Stroke patients’ functions in personal activities of daily living in relation to sleep and socio-demographic and clinical variables in the acute phase after first-time stroke and at six months of follow-up. J Clin Nurs. Jul 2012;21(13-14):1886–95. doi:10.1111/j.1365-2702.2011.04014.x

28. Williams-Cooke C, Watts E, Bonnett J, Alshehri M, Siengsukon C. Association Between Sleep Duration and Functional Disability in Inpatient Stroke Rehabilitation: A Pilot Observational Study. Archives of Rehabilitation Research and Clinical Translation. 2021;3(3)doi:10.1016/j.arrct.2021.100150

29. Kim WH, Yoo YH, Lim JY, et al. Objective and subjective sleep problems and quality of life of rehabilitation in patients with mild to moderate stroke. Top Stroke Rehabil. Apr 2020;27(3):199–207. doi:10.1080/10749357.2019.1673591

30. Fleming MK, Smejka T, Henderson Slater D, et al. Sleep Disruption After Brain Injury Is Associated With Worse Motor Outcomes and Slower Functional Recovery. Neurorehabil Neural Repair. Jun 7 2020:1545968320929669. doi:10.1177/1545968320929669

31. Selves C, Stoquart G, Lejeune T. Gait rehabilitation after stroke: review of the evidence of predictors, clinical outcomes and timing for interventions. Acta Neurol Belg. Mar 12 2020;doi:10.1007/s13760-020-01320-7

32. Duarte E, Marco E, Muniesa JM, et al. Trunk control test as a functional predictor in stroke patients. J Rehabil Med. Nov 2002;34(6):267–72.

33. Liao WL, Chang CW, Sung PY, Hsu WN, Lai MW, Tsai SW. The Berg Balance Scale at Admission Can Predict Community Ambulation at Discharge in Patients with Stroke. Medicina (Kaunas). May 31 2021;57(6)doi:10.3390/medicina57060556

34. Harari Y, O’Brien MK, Lieber RL, Jayaraman A. Inpatient stroke rehabilitation: prediction of clinical outcomes using a machine-learning approach. J Neuroeng Rehabil. Jun 10 2020;17(1):71. doi:10.1186/s12984-020-00704-3

35. Siengsukon C, Boyd LA. Sleep enhances off-line spatial and temporal motor learning after stroke. Neurorehabil Neural Repair. May 2009;23(4):327–35. doi:10.1177/1545968308326631

36. Siengsukon CF, Boyd LA. Sleep to learn after stroke: implicit and explicit off-line motor learning. Neurosci Lett. Feb 13 2009;451(1):1–5. doi:10.1016/j.neulet.2008.12.040

37. Siengsukon CF, Boyd LA. Sleep enhances implicit motor skill learning in individuals poststroke. Top Stroke Rehabil. Jan-Feb 2008;15(1):1–12. doi:10.1310/tsr1501-1

38. Stewart NH, Arora VM. Sleep in Hospitalized Older Adults. Sleep Med Clin. Mar 2018;13(1):127–135. doi:10.1016/j.jsmc.2017.09.012

